# A realist approach to implementation fidelity in a mixed-method evaluation of electronic decision support systems to improve the quality of antenatal care in Nepal

**DOI:** 10.1101/2024.05.07.24306757

**Authors:** Emma Radovich, Sulata Karki, Seema Das, Rajani Shakya, Ona L. McCarthy, Abha Shrestha, Clara Calvert, Oona M. R. Campbell, Loveday Penn-Kekana

## Abstract

**Background:** Understanding implementation fidelity, or adherence to the intervention-as-intended, is essential to interpreting the results of evaluations. In this paper, we propose a longitudinal, explanatory approach to implementation fidelity through a realist evaluation lens. We apply this approach to a mixed-method assessment of implementation fidelity to an electronic decision support system intervention to improve the quality of antenatal care in Nepal.

**Methods:** The tablet-based electronic decision support system was implemented in 19 primary care facilities in Nepal. As part of the project’s process evaluation, we used four data sources – monitoring visit checklists and fieldnotes, software backend data, and longitudinal case studies in four facilities – to examine three components of fidelity: use at the point of care, use for all antenatal visits, and quality of data entry. Quantitative data were analysed descriptively. Qualitative data were analysed thematically using template analysis to examine descriptive findings across the three fidelity components and later to develop and reflect on the causal mechanisms. Findings were synthesised, drawing on Normalization Process Theory, to understand the processes driving the different patterns of fidelity observed.

**Results:** Fidelity to point-of-care use declined over time with healthcare providers often entering data after antenatal visits had ended because providers understood the intervention as primarily about recordkeeping rather than decision support. Even in facilities with higher fidelity to point-of-care use, software decision-support prompts were largely ignored. Low antenatal client caseloads and the suggestion by fieldworkers to practice back-entering data from previous antenatal visits undermined understanding of the intervention’s purpose for decision support.

**Conclusions:** Our assessment explains how and why patterns of implementation fidelity occurred, yielding more nuanced understanding of the project evaluation’s null result that moves beyond intervention vs implementation failure. Our findings demonstrate the importance of discussing intervention theory in terms fieldworkers and participants understand so as not to undermine fidelity.

## Introduction

This paper reports on the conceptual approach and empirical findings of a mixed-method assessment of implementation fidelity within an evaluation of a digital health intervention to improve the quality of antenatal care (ANC) in Nepal. We propose a longitudinal, explanatory approach to implementation fidelity, drawing on realist evaluation, to examine how fidelity unfolded and why. In doing so we contribute to evidence about the realities of intervention implementation in maternal health and deepen understanding of how efforts to ensure ANC quality can be improved(1).

Evaluations are frequently concerned with implementation fidelity, or the consistency with which the intervention was implemented as intended(2–5). When an intervention is ineffective, process evaluations are often tasked with determining whether it was because it does not work (intervention failure) or because it was not implemented as intended (implementation failure), seeing this as an internal validity question about whether the outcome evaluation was a valid test of the intervention theory(2,5–7). However, this neat, suggested divide between intervention and implementation failure stems from a positivist view of evaluation. Fidelity assessment in this mode is often about trying to discern the so-called ‘true effect’ of the intervention(8), and lack of fidelity is frequently blamed for why studies of seemingly the same intervention led to different results in different contexts(4,8).

Our ideas about what implementation fidelity is and how it should be examined are informed by a realist approach to evaluation. Realist evaluation, and more recent perspectives on the evaluation of complex interventions, emphasize explanation – how, for whom and under what circumstances do interventions work(2,9,10). Evaluations from a realist perspective understand interventions as shaped by their contexts and that these contextually-dependent adaptations may trigger mechanisms leading to desired outcomes or, conversely, unintended consequences(11,12). We argue a realist evaluation approach to fidelity moves beyond an inward-looking emphasis on internal validity to develop more outward-looking explanation of how and why the intervention-as-intended interacted with its context to produce the process of implementation—and the resulting outcomes—observed. Assessing fidelity requires examining how closely (or not) the implementation process followed what the intervention designers had hoped would happen. A realist understanding of fidelity would see that this is not a binary (fidelity vs implementation failure) but has degrees of consistency and is a process that can change over time and unfold in a non-linear fashion(7,13– 15).

We apply these ideas to the Mobile health Integrated Rural Antenatal care (mIRA) implementation research project in Nepal, which aimed to improve the quality of ANC using electronic decision support systems (EDSS) introduced at primary care facilities. EDSS are information systems, often delivered through computers or tablets, that integrate clinical and demographic data to support healthcare providers’ decision-making and improve adherence to guidelines via checklists, alerts or information provided at the point of care(16,17). The mIRA project evaluated two tablet-based EDSS: the newly developed mIRA EDSS(18) and the World Health Organization (WHO) digital ANC module(19), using a two-phase outcome assessment, comparing quality scores pre- and post-EDSS implementation from observations of ANC consultations, alongside a robust process evaluation(20,21). The mIRA project evaluation results are reported in detail elsewhere(21), but for the most part, the EDSS intervention did not improve quality of care outcomes.

All interventions embody assumptions about how a programme reaches its anticipated outcomes(2,11). For the EDSS to improve quality of care, we assumed providers would incorporate the EDSS into their workflow so that they could input data into the tablets, respond to its prompts, and make the desired changes to their clinical practice. Project investigators identified three essential components of using the intervention as intended: 1) providers should fill-in the EDSS during consultations with pregnant women; 2) providers should use the EDSS for all ANC visits; and 3) providers should enter sufficient data in the EDSS so that the algorithms generate recommendations and reminders. The three components were based on the idea that point-of-care support improves adherence to guidelines through ‘prompts to action’ reminding healthcare providers of what they should do in their clinical practice at the time and location of decision making(22,23). Use at the point of care (component 1) was a vital component in the intervention theory for how the EDSS would improve ANC quality. For example, the EDSS included an alert to perform a dipstick test if the results of a urine protein test were not recorded for each ANC consultation. If the pregnant woman had finished her ANC consultation and left the clinic, then the provider entering information into the EDSS later would be unable to follow this prompt and perform the test. For components 2 and 3, the mIRA and WHO EDSS were understood to work best if used at every ANC visit to enable the diagnostic algorithms for longitudinal care throughout the pregnancy. Use for all ANC visits would also minimize the need to back-enter data from previous patient contacts, a factor known to hinder EDSS uptake and effectiveness in other settings(16).

This study, as part of the mIRA project’s process evaluation, offers a realist assessment of implementation fidelity to the mIRA EDSS and WHO EDSS intervention-as-intended in Nepal. We aim to give a rationale for widening assessment of implementation fidelity and to demonstrate how we operationalised and analysed these ideas. We do this by 1) describing implementation fidelity over time and between facilities implementing the two EDSS, using the three components of EDSS intervention-as-intended, described above, and 2) developing explanations for how implementation contexts shaped mechanisms that led to observed differences in fidelity.

## Methods

### Intervention and setting

The mIRA project took place in four predominantly rural districts in Bagmati Province, Nepal (Kavrepalanchok, Sindhupalchowk, Sindhuli and Dolakha) between April and December 2022. Twenty facilities—government Health Posts and Primary Health Care Centers and non-governmental Dhulikhel Hospital Outreach Centers—were selected to receive tablets with the EDSS software installed(20). Facilities were paired by type and reported ANC client volume for the previous year and then randomly allocated to receive either the mIRA or the WHO EDSS. Following allocation, one facility assigned to the WHO EDSS arm was discovered to have extremely low client volume (<5 ANC cases/year) and was dropped, without replacement, from the project.

Each facility received a tablet with the allocated EDSS software installed and glucometers to facilitate performance of oral glucose tolerance tests when prompted by the EDSS, as this equipment was not normally present in the facilities. One Auxiliary Nurse Midwife (ANM) from each facility working in ANC was selected by the local municipality to attend a three-day training workshop hosted by Dhulikhel Hospital on either the mIRA or WHO EDSS. The workshops consisted of training on the purpose of the EDSS and hands-on practice entering data in the EDSS. Participants additionally received training in administering oral glucose tolerance tests.

The trained ANMs were then supported in using their allocated EDSS by an onsite fieldworker during a monthlong lead-in period, during which other ANMs (who did not attend the training) could also receive instruction in EDSS use. ANMs were encouraged to use the EDSS during consultations with real ANC clients. However, due to low client volume in some facilities, fieldworkers would sometimes encourage ANMs to practice using the EDSS with dummy data or by snapping photos of a pregnant woman’s paper ANC card and entering data from past ANC visits. ANMs were expected to continue to complete paper-based records (ANC cards and ANC registers) alongside the EDSS, during the project.

### Study design and data collection

The implementation fidelity study used a mixed-method convergent design where quantitative and qualitative data were collected simultaneously(24). The intent was to offer a more complete understanding of implementation in all 19 facilities, drawing on quantitative and qualitative data from in-person monitoring visits, quantitative information captured in the backend data of the EDSS software, and repeat, unstructured observations and in-depth interviews conducted in four case study facilities(20).

Following the supported lead-in period, project fieldworkers visited facilities with a structured checklist to assess functioning of the tablet and EDSS software, whether the EDSS was observed in use on the day and the number of entries on the facility’s ANC register for the previous day (or the date of the last register entry). Fieldworkers conducting the monitoring visits were trained by the project’s anthropologist (LPK) to additionally document, via free-form notes at the end of the structured checklist, how the ANMs described using the EDSS and problems encountered with the tablets or software. The fieldnotes had a dual purpose: firstly, to enable project staff to identify and reconcile problems, for example providing replacement memory cards for tablets. Secondly, the fieldnotes captured whether and how ANMs were using the EDSS based on the fieldworkers’ observations and informal conversations with facility staff. Fieldworkers conducted four monitoring visits at intervals of 1-2 months at each facility during the implementation period; however, weather-related issues made it impossible to conduct two (of the four) in-person visits at one facility and one visit at another facility.

We extracted data logged via the EDSS software. The backend data included the facility identification code, the date and number of visit entries logged (to compare to the number of ANC register entries), and values recorded for a selection of non-mandatory fields. Most software fields were mandatory, and those that were non-mandatory were different in each EDSS. We selected three non-mandatory fields in each EDSS, identifying those likely to be relevant in all ANC visits and relevant to the quality outcome measures(20). The selected fields for the WHO EDSS were: 1) counselling on next visit schedule, 2) was pallor assessed, and 3) was oedema assessed; selected non-mandatory fields for the mIRA EDSS were: 1) any current pregnancy complaints, 2) was urine protein done, and 3) was haemoglobin test done. Several mIRA EDSS facilities experienced software-related difficulties with saving and syncing data to the server, meaning entries were saved locally on the tablet but not visible in the backend data. Software issues were resolved and EDSS entries synced, part-way through implementation for six of the 10 mIRA-allocated facilities. The four mIRA facilities with ongoing syncing issues are missing an unknown number of visit entries in the backend data; results from these facilities are presented separately.

We conducted longitudinal case studies in four purposively selected facilities, two each implementing the mIRA and WHO EDSS, to explore changes in ANC provision and facility operations over the course of the intervention(21,25). Two researchers (SK and SD) conducted repeat observations, formal interviews, and findings validation workshops with facility staff. The researchers documented daily their observations and reflections and transcribed the formal interviews; we also took extensive notes during regular team discussions throughout the data collection period, responding to emerging findings and developing and testing hypotheses(25). For the purposes of this analysis, we examined evidence in the notes and transcripts on how and why ANMs used the EDSS in the way that they did, based on the three components of fidelity.

Data from all sources were collected simultaneously during the four rounds of in-person monitoring visits and longitudinal case study observations. Initial analyses for this study began during the final phase of data collection for the fourth in-person monitoring visit.

### Definitions

Carroll and colleagues’ conceptual framework for implementation fidelity, where components are evaluated to understand “whether the result of the implementation process is an effective realisation of the intervention as planned by its designers”(4) offered a useful guide for defining components of fidelity in this study(26). We mapped our three components to elements of Carroll and colleagues’ domain of adherence(4) (Table 1). While the framework considers quality of delivery as a moderator of adherence, we considered quality, or how closely EDSS use approached the theoretical ideal of complete data entry enabling full software functionality, as a discrete aspect of fidelity due to the importance of this component in our intervention theory.

**Table 1.** Components of implementation fidelity and their operationalisation in the study.

| EDSS components of fidelity | How we mapped to Carroll and colleagues' concepts in the conceptual framework for implementation fidelity | Operationalisation of the concept across the data sources |
| --- | --- | --- |
| 1. Providers should fill-in the EDSS during consultations with pregnant women. | **Content:**<br>Timeliness of use of the EDSS, that is at the point-of-care, was considered a crucial 'active ingredient' of how the intervention sought to deliver a change in provider practice. | <u>Monitoring visit checklist:</u> Proportion of ANC consultations with observed use of EDSS, and proportion of facilities with tablet 'ready to use'.<br><u>Monitoring fieldnotes and longitudinal case studies:</u><br>Examining point-of-care use vs recordkeeping after. |
| 2. Providers should use the EDSS for all ANC visits. | **Frequency (or coverage):**<br>Consistency of use of the EDSS for all ANC visits links closely to the elements that comprise 'dose' in the framework: frequency, coverage and duration. We examine duration via our longitudinal approach using repeated measures. | <u>Monitoring visit checklist and backend data:</u><br>Proportion of register entries logged in EDSS per monitoring visit.<br><u>Monitoring fieldnotes and longitudinal case studies:</u><br>Examining use for every visit vs rationalizing when to use or not use. |
| 3. Providers should enter sufficient data in the EDSS so that the algorithms can generate recommendations and reminders. | **Quality:**<br>Many fields in both EDSS were marked mandatory (visit entries could not be saved without a value entered), essentially ensuring this component's minimum fidelity. So, we examined whether quality of use of the EDSS went beyond what was mandatory to the scope of EDSS functionality that was enabled. | <u>Backend data:</u><br>Proportion of selected non-mandatory fields completed out of total number of EDSS entries created, where to be considered complete the field could have any option selected or be marked as 'not done'. Fields with nothing entered were considered incomplete.<br><u>Monitoring fieldnotes and longitudinal case studies:</u><br>Examining how providers made use of EDSS functionality or discrepancies between performing tasks/actions and recording them. |

For each in-person monitoring visit, we assessed whether the tablet was ‘ready to use’: the tablet was available, reported functional, reported at least 30% charged (based on estimated battery life needed to record an ANC visit and synchronize files to the server) and connected to either the mobile or internet network. Tablets that were reported unavailable, non-functional, insufficiently charged or with no network connection were considered not ready to use. Tablets insufficiently charged were considered ready to use if there was a functional charging cord available and electricity available at the facility.

### Data analysis

We used a modified parallel-databases approach in which the quantitative and qualitative data were analysed independently, using the data to examine aspects of the three fidelity components(24). Findings from the quantitative and qualitative analyses were brought together during the integration and interpretation stage. Integration of the quantitative and qualitative results involved identifying content areas represented in the multiple datasets and creating a joint display matrix to merge the results for each facility. Descriptive statistics for the related quantitative variables and qualitative data relating to the three core fidelity domains of content, frequency, and quality of EDSS use were arrayed in the matrix. Quantitative and qualitative findings in the matrix were given equal emphasis.

### Quantitative analysis

Quantitative data from the monitoring visit checklists and the EDSS backend were analysed descriptively. Analyses were conducted in Stata/SE V.16 (StataCorp, College Station, Texas, USA). No tests of statistical significance were performed as the study was not designed to test for differences in outcomes and the sample sizes were not powered to do so(20).

Analysis of the EDSS backend data included all entries created with a visit date during the implementation period: from the approximate end of the lead-in period through the end of supported implementation for the six Nepali months of Jestha to Kartik 2079 (corresponding to Gregorian calendar dates 15 May to 16 November 2022). No entries were missing visit dates; this was a mandatory field in mIRA EDSS or was created automatically in WHO EDSS. A small number of entries (<5%) were marked as “demo” or test entries and were dropped before the analysis.

Completeness of backend data was assessed for the mIRA and WHO EDSS separately as a single cross-sectional measure of the proportion of each non-mandatory field completed out of the total number of EDSS entries. Blank variable fields (nothing recorded) were considered incomplete. Anything entered in the field, including ‘not done’, were considered complete.

For the comparison between the previous day’s number of entries on the ANC register and the number of entries logged in the EDSS, we tallied the number of records saved in the backend data for each facility for the designated ANC register date for each of the four rounds of monitoring visits. Some dates on the ANC register included zero entries (no ANC clients attended on that day). Agreement was calculated as an exact match in the number of entries, including zero entries, recorded in the ANC register and the number of visits recorded in the EDSS for the same date. Analysis comparing numbers of register and backend entries for each facility was done in Excel.

### Qualitative analysis

Fieldnotes and interview transcripts from the longitudinal case studies were analysed thematically by SK, SD and LPK (detailed methods for the longitudinal case studies published separately(25)). For the purposes of this study, themes arising from the longitudinal case studies analysis that related to the three fidelity components were extracted and included here.

Monitoring visit fieldnotes were assembled for each facility. Template analysis of the fieldnotes was based on an initial codebook with *a priori* themes based around the three fidelity components. Template analysis is a flexible approach to thematic analysis and tends to define themes from a mix of *a priori* interests and initial engagement with the data before applying the codebook to the full dataset(27). The initial codebook was developed and fieldnotes were analysed by ER; the monitoring fieldnotes for two facilities were also analysed independently by two research team members (SK and LPK). Themes were compared and refined in a reflexive process intended to increase rigour and deeper analysis. The monitoring fieldnotes were coded and analysed using NVivo.

The data were initially coded descriptively, that is, the codes addressed the initial research aim of describing patterns in the components of fidelity. The research aim of explaining how different patterns arose, shifted the analysis to creating consolidated codes, and re-reading the data to check the codes’ interpretive validity, to develop themes of causal explanation(28). We focused analysis on the contextual factors and mechanisms shaping the process of implementation fidelity(29), rather than the overall outcome of the project’s evaluation comparing quality of care measures before/after EDSS implementation, which is reported elsewhere(21).

### Integration of results

Analyses were completed concurrently and iteratively, moving between quantitative and qualitative analyses to develop and interpret how patterns of implementation fidelity were shaped by context and mechanisms over time, drawing on theories of implementation(29–31). The analysis was guided by Normalization Process Theory, a theory of action organised around four key constructs (coherence, cognitive participation, collective action and reflexive monitoring) that describe the different types of work needed to take something new (the EDSS intervention) and make it part of routine practice(30). The constructs and underlying generative mechanisms of Normalization Process Theory interact, and implementation fidelity was shaped by mechanisms operating in unique contexts. Using May and colleagues’ recent guidance on linking Normalization Process Theory constructs to context-mechanism-outcome configurations(29), we examined how contexts, including organising structures and group processes, affected the dynamics of EDSS implementation. We drew from work conceptualising interventions as events within systems(15) to think about how mechanisms operated within the plasticity of the intervention (the extent to which users could modify intervention components) and how tightly or loosely the intervention was coupled with its implementation context (the level of negotiation users had in interacting with the intervention and the extent of work demanded to adapt the intervention to their contexts)(31). Monitoring fieldnotes were coded deductively against the four Normalization Process Theory constructs with further themes arising inductively, and these were compared and integrated with findings from the quantitative analyses. Preliminary explanations were reviewed by senior co-authors (LPK and OMRC) in regular meetings to enhance the reliability of findings. Finally, the research team reflected on the validity of the explanations, their plausibility and explanatory power(28).

### Ethical approval

The mIRA project, in which this analysis was part, received ethical approval from Kathmandu University School of Medical Sciences’ Institutional Research Committee (ref: IRC-KUSMS 25/22), Nepal Health Research Council’s Ethical Review Board (ref: 2695) and the London School of Hygiene & Tropical Medicine’s Intervention Research Ethics Committee (ref: 25094-1).

## Results

We firstly present descriptive findings across the three components of fidelity, followed by explanations of the mechanisms operating within contexts to shape implementation fidelity.

Table 2 shows results from the four in-person monitoring visits. The length of time between monitoring visits increased over the duration of the project, from between 18-38 days between the first two monitoring visits to between 32-105 days for the final two monitoring visits.

**Table 2.** Results of in-person monitoring visits to all facilities.

|  | WHO facilities (n=9) | mIRA facilities (n=10) |
| --- | --- | --- |
| **Monitoring visit 1 (20 May – 2 Jun 2022)** |  |  |
| Number of facilities with monitoring visit | 9 | 10 |
| Proportion of facilities with tablet ready to use | 100% | 80% |
| Mean number of days since last sync with server | 0.0 (SD*: 0.0) | 2.2 (SD: 2.3) |
| Number of facilities with ANC consultation observed | 3 | 2 |
| Proportion of ANC consultations with observed use of EDSS | 100% | 100% |
| **Monitoring visit 2 (19 Jun – 28 Jun 2022)** |  |  |
| Number of facilities with monitoring visit | 8 | 10 |
| Range in number of days between monitoring visit 1 and 2 | 18 – 32 | 23 – 38 |
| Proportion of facilities with tablet ready to use | 100% | 80% |
| Mean number of days since last sync with server | 0.1 (SD: 0.35) | 5.0 (SD: 9.8) |
| Number of facilities with ANC consultation observed | 3 | 3 |
| Proportion of ANC consultations with observed use of EDSS | 100% | 100% |
| **Monitoring visit 3 (18 Jul – 18 Aug 2022)** |  |  |
| Number of facilities with monitoring visit | 8 | 9 |
| Range in number of days between monitoring visit 2 and 3 | 21 – 56 | 21 – 49 |
| Proportion of facilities with tablet ready to use | 100% | 78% |
| Mean number of days since last sync with server | 0.0 (SD: 0.0) | 2.3 (SD: 2.7) |
| Number of facilities with ANC consultation observed | 4 | 4 |
| Proportion of ANC consultations with observed use of EDSS | 75% | 50% |
| **Monitoring visit 4 (2 Sep – 16 Nov 2022)** |  |  |
| Number of facilities with monitoring visit | 9 | 10 |
| Range in number of days between monitoring visit 3 and 4 | 35 – 98 | 32 – 105 |
| Proportion of facilities with tablet ready to use | 100% | 100% |
| Mean number of days since last sync with server | 0.1 (SD: 0.3) | 0.8 (SD: 1.0) |
| Number of facilities with ANC consultation observed | 5 | 4 |
| Proportion of ANC consultations with observed use of EDSS | 80% | 50% |

\*SD = standard deviation

### Content: point-of-care use

Across the four monitoring visits, all WHO facilities and nearly all mIRA facilities had the tablet ‘ready to use’ (available, functional and sufficiently charged) (Table 2). Few facilities (20-55% of facilities in each of the four visits) had an ANC consultation during the monitoring visit. Among facilities with an ANC consultation observed, all were observed to use the EDSS at the point of care during the first two monitoring visits. Point-of-care use reduced to 50-80% for consultations observed in the third and fourth monitoring visits (Table 2).

Results from the monitoring visit fieldnotes and longitudinal case studies suggested ANMs used the EDSS primarily for recordkeeping, often entering data after the consultation, rather than as a point-of-care tool (Table 3). In many instances, providers took a photograph of the pregnant woman’s handheld ANC card before she left the facility and entered the information into the EDSS later:

**Table 3.**
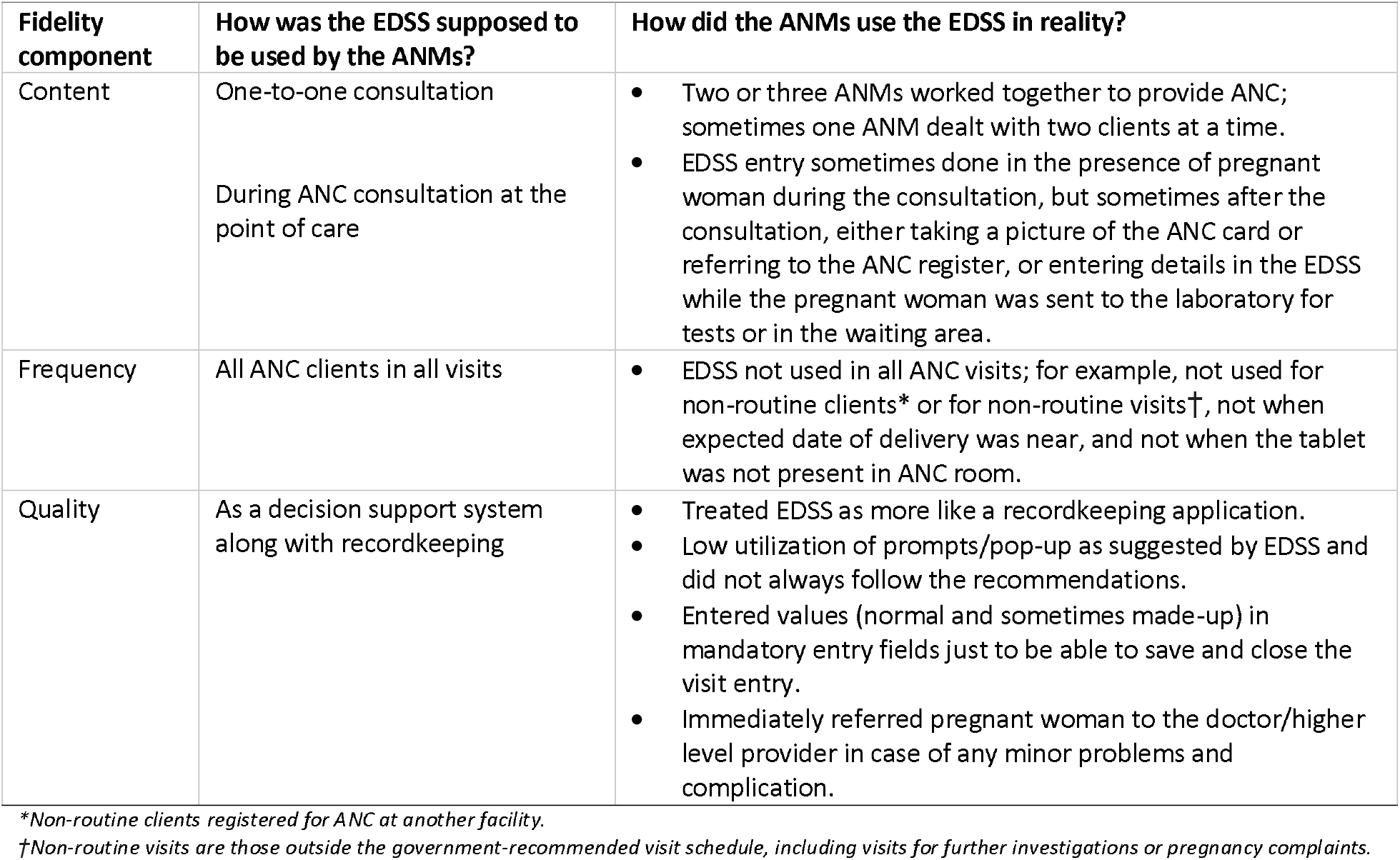
Synthesis of findings from the longitudinal case studies in four facilities.

| **Fidelity component** | **How was the EDSS supposed to be used by the ANMs?** | **How did the ANMs use the EDSS in reality?** |
| --- | --- | --- |
| Content | One-to-one consultation<br>During ANC consultation at the point of care | Two or three ANMs worked together to provide ANC; sometimes one ANM dealt with two clients at a time.<br>EDSS entry sometimes done in the presence of pregnant woman during the consultation, but sometimes after the consultation, either taking a picture of the ANC card or referring to the ANC register, or entering details in the EDSS while the pregnant woman was sent to the laboratory for tests or in the waiting area. |
| Frequency | All ANC clients in all visits | EDSS not used in all ANC visits; for example, not used for non-routine clients* or for non-routine visits†, not when expected date of delivery was near, and not when the tablet was not present in ANC room. |
| Quality | As a decision support system along with recordkeeping | Treated EDSS as more like a recordkeeping application.<br>Low utilization of prompts/pop-up as suggested by EDSS and did not always follow the recommendations.<br>Entered values (normal and sometimes made-up) in mandatory entry fields just to be able to save and close the visit entry.<br>Immediately referred pregnant woman to the doctor/higher level provider in case of any minor problems and complication. |

\*Non-routine clients registered for ANC at another facility.
†Non-routine visits are those outside the government-recommended visit schedule, including visits for further investigations or pregnancy complaints.

> *“Three ANMs are responsible for ANC check up and all of them uses the mIRA application. I asked one of the ANM staff, who was present in the ANC room, whether they have used mIRA application at the time of ANC consultation, then she replied that they don’t use it when they are busy with high patient flow. They just click the photos of ANC card and filled it later in the application*.*”* (*Monitoring fieldnotes, 2*^*nd*^ *monitoring visit, mIRA facility*)

Busy periods with multiple pregnant women attending the facility, including on designated ANC days, were frequently mentioned as reasons why the EDSS was not used or why data was entered later. ANMs working alone struggled to incorporate the EDSS into the consultation, and point-of-care use was achieved when the ANMs worked together to divide the tasks of documentation and conducting the consultation:

> *“Mostly the trained ANM is using the WHO EDSS. We also observe untrained ANM using the EDSS with the help of trained ANM during the consultation. Today* (*Thursday*) *is the ANC day, so day is a bit busy. ANMs* (*3*) *are working together and they are simultaneously using EDSS and recording as per the variables in the ANC card*.*”* (*Monitoring fieldnotes, 1*^*st*^ *monitoring visit, WHO facility*)

### Frequency: use for all visits

Table 4 shows the comparison in the number of entries recorded in the ANC register for a specific date and the number of entries logged in the EDSS for the same date for four rounds of monitoring visits. Among all facilities, agreement in number of entries (which included zero entries) varied between 28% to 65% in the four monitoring visits, without a clear pattern over time. In each round of monitoring visits, a substantial proportion (24-58%) of dates saw fewer EDSS entries recorded compared to entries in the ANC register for the same day. Agreement in number of EDSS and register entries was lower among mIRA facilities than WHO facilities for all monitoring visits, except for the mIRA facilities with syncing issues in monitoring visit three. For WHO facilities, there was a small reduction in dates with agreement in numbers of entries documented (78% to 56%) and an increased proportion of dates with fewer entries in the EDSS compared to the register (11% to 44%) from the first to the fourth monitoring visit.

**Table 4.** Comparison of number of ANC visit entries recorded in the paper-based ANC register and the EDSS software backend data across four rounds of monitoring visits.

|  | WHO facilities (n=9) | mIRA facilities without syncing issues (n=6) | mIRA facilities with syncing issues (n=4) | TOTAL (n=19) |
| --- | --- | --- | --- | --- |
|  | n (%) | n (%) | n (%) | n (%) |
| **Monitoring visit 1** |  |  |  |  |
| Number of facilities with monitoring visit | 9 | 6 | 4 | 19 |
| Agreement in number of EDSS and register entries* | 7 (78%) | 2 (33%) | 1 (25%) | 10 (53%) |
| Zero entries in register and in EDSS | 1 (11%) | 1 (17%) | 0% | 2 (11%) |
| Fewer entries in EDSS than in register | 1 (11%) | 4 (67%) | 2 (50%) | 37 (7%) |
| **Monitoring visit 2** |  |  |  |  |
| Number of facilities with monitoring visit | 8 | 6 | 4 | 18 |
| Agreement in number of EDSS and register entries* | 5 (63%) | 0 (0%) | 0 (0%) | 5 (28%) |
| Zero entries in register and in EDSS | 3 (38%) | 0 (0%) | 0 (0%) | 3 (17%) |
| Fewer entries in EDSS than in register | 2 (25%) | 5 (83%) | 3 (75%) | 10 (56%) |
| **Monitoring visit 3** |  |  |  |  |
| Number of facilities with monitoring visit | 8 | 6 | 3 | 17 |
| Agreement in number of EDSS and register entries* | 5 (63%) | 3 (50%) | 3 (100%) | 11 (65%) |
| Zero entries in register and in EDSS | 1 (13%) | 2 (33%) | 1 (33%) | 4 (24%) |
| Fewer entries in EDSS than in register | 2 (25%) | 2 (33%) | 0 (0%) | 4 (24%) |
| **Monitoring visit 4** |  |  |  |  |
| Number of facilities with monitoring visit | 9 | 6 | 4 | 19 |
| Agreement in number of EDSS and register entries* | 5 (56%) | 2 (33%) | 1 (25%) | 8 (42%) |
| Zero entries in register and in EDSS | 2 (22%) | 1 (17%) | 0 (0%) | 3 (16%) |
| Fewer entries in EDSS than in register | 4 (44%) | 4 (67%) | 3 (75%) | 11 (58%) |

\*Includes both equal number of entries in EDSS as register and zero entries in register and in EDSS.

The EDSS was rarely used when pregnant women visited the facility for additional investigations, such as ultrasound (USG) scans, or when they were not regular clients registered at the facility (Table 3). This aligned use of the EDSS with existing practices on who and what gets documented in paper ANC records:

> *“The tablet is being used for those cases which is a regular case at the facility but not those come for USG and lab investigation*.*”* (*Monitoring fieldnotes, 4*^*th*^ *monitoring visit, mIRA facility*)

> *“ANC consultation from another ward details are only entered in [EDSS] if ANC card and test reports are brought by the patient, otherwise data is not entered in [EDSS]. Also, their registration is not done in ANC register*.*”* (*Monitoring fieldnotes, 1*^*st*^ *monitoring visit, WHO facility*)

For ANC visits that would have been documented in the ANC register, as in Table 4, EDSS entry was often limited to the ANM who had received workshop training in using the tablet and software. ANMs described how the availability of the trained ANM, whether due to leave or assignment to non-ANC duties, shaped use of the EDSS and meant that some ANC visits did not get entered:

> *“In my previous monitoring visit the trained ANM was on leave for one month, and only 1 ANM was there and there was no tablet in health facility. But this time the trained ANM was there with tablet. The trained ANM told me that the tablet was not being used when she was on leave for a month. Since other ANMs does [sic] not show interest in using the tablet with EDSS, she did not [leave] the tablet at health facility. And tablet hasn’t been used by anyone else for that period of time. She told me that she is the only one in the health facility who uses the tablet. When I asked her ‘why does any other staffs do not use the tablet?’ She answered that other staffs doesn’t show any interest on using the app and also believed that only trained ANM supposed to use the tablet*.*”* (*Monitoring fieldnotes, 3*^*rd*^ *monitoring visit, mIRA facility*)

### Quality: scope of EDSS functionality enabled

Table 5 shows the proportion of selected non-mandatory fields completed out of the total number of EDSS entries created; completeness was nearly universal (>99%) for all three variables in the WHO EDSS. Completeness for the selected three non-mandatory fields in the mIRA EDSS ranged from 40% to 90%, with only one-third of entries having all three fields completed. The mIRA EDSS non-mandatory fields of urine protein test and haemoglobin test were the least completed variables assessed. These tests require equipment that may not have been available at the facility during the ANC visit, so it was possible to select ‘not done’ in the EDSS and give stock-out of test kits as the reason.

**Table 5.** Completeness of EDSS backend data for all visit entries dated during the implementation period (15 May – 16 Nov 2022) for the mIRA EDSS and WHO EDSS.

|  | Non-mandatory field | Value recorded in EDSS | Proportion of entries complete |
| --- | --- | --- | --- |
| **mIRA EDSS**<br>n=848 entries | Current pregnancy complaints | Any option selected/no complaints reported [vs nothing entered] | 90% |
|  | Urine protein test | Done/not done [vs nothing entered] | 40% |
|  | Haemoglobin test | Done/not done [vs nothing entered] | 40% |
|  | All three variables |  | 34% |
| **WHO EDSS**<br>n=987 entries | Visit schedule counselling | Done/not done [vs nothing entered] | 100% |
|  | Pallor assessed | Yes/no [vs nothing entered] | 100% |
|  | Oedema present | Yes present/no [vs nothing entered] | 100% |
|  | All three variables |  | 99% |

The monitoring fieldnotes and longitudinal case studies suggested ANMs rarely used the prompts or recommendations from the EDSS and in some cases recorded care components without performing them, often out of frustration with trying to fill in all the mandatory fields to save the EDSS entry (Table 3). ANMs described the EDSS as something for recordkeeping, though some noted that the entry form could serve as a helpful reminder to do things. But custom prompts and pop-ups (“toasters”) were ignored, and ANMs often did not make use of some of the software’s functionality, such as the auto-calculation of gestational age:

> *“She was calculating week of gestation in rough page and copied in ANC card. Completed consultation and filing in ANC card then used WHO app for ANC as this was the client for 4*^*th*^ *visit to health facility in 9 mth. Trained ANM used the app and was providing counselling but did not check for the toasters in the application*.*”* (*Monitoring fieldnotes, 1*^*st*^ *monitoring visit, WHO facility*)

### Explaining patterns of implementation fidelity

Table 6 outlines how Normalization Process Theory constructs appeared in our examination of implementation fidelity. The generative mechanisms underlying these constructs shaped how implementation fidelity unfolded in the different facilities.

**Table 6.** Normalization Process Theory constructs and interpretation of how these mechanisms appeared in our assessment of EDSS implementation fidelity.

| **Normalization Process Theory construct** | **How the constructs and their related components appeared in our study** |
| --- | --- |
| *Coherence* – sense-making work: understanding what was different about the EDSS intervention, understanding its aims, understanding what the individual's responsibilities/tasks were in relation to the EDSS and the value/benefits of these activities. | *Differentiation:*<br>Participating ANMs understood the EDSS as another recordkeeping practice and did not see it as a tool to support decision making or to change how they provided care during ANC visits.<br>*Internalization:*<br>Few ANMs spoke of its potential usefulness as a reminder, particularly for counselling topics. |
| *Cognitive participation* – relational work: whether key participants (e.g., the trained ANM) drove the new practice forward, whether ANMs believed it was right for them to be involved in the EDSS intervention and how they rethought group relationships. | *Initiation and legitimization:*<br>Enthusiasm or interest in learning to use the EDSS did not always align with the ANM selected to receive training, and some ANMs who did not receive EDSS training did not view using the EDSS as a legitimate part of their role.<br>*Organisational logic and enrolment:*<br>Many facility staff saw the EDSS intervention as a time-limited research study that was tangential and duplicative to the government-allocated work of documentation in ANC cards and monthly reporting. |
| *Collective action* – operational work: how ANMs operationalised use of the tablet during ANC visits and the resources needed for this. | *Interactional workability:*<br>Teams of ANMs worked together to incorporate EDSS data entry into the process of ANC consultations and other forms of documentation.<br>*Skill-set workability:*<br>The EDSS software was not always intuitive to use, and low ANC caseloads meant ANMs had few opportunities to practice and consolidate skills in using the software. ANMs developed workarounds to adapt the EDSS to fit with paper-based recordkeeping practices, such as entering information about twin pregnancies using '/' marks in relevant variable fields in both the paper records and EDSS. |
| *Reflexive monitoring* – appraisal work: ANMs reflecting on whether participation in the new tasks of the EDSS intervention was working, including how the intervention was refined to make it workable in practice, how it affected them and how useful it was. | *Individual appraisal:*<br>Any potential benefits in decision support could not overcome the increased workload that accompanied increased data entry required by the EDSS. The EDSS was blamed for increasing the length of consultations when ANC clients complained about long wait times.<br>*Reconfiguration:*<br>ANMs redefined prompted procedures to fit within existing practices, such as only doing oral glucose tolerance tests (provided by the mIRA project intervention and prompted by the EDSS as the preferred action) if the random blood sugar test result was high. |

The EDSS was almost universally understood as a recordkeeping task (Table 6), which shaped how the EDSS was used: taking of photos of ANC cards for later data entry, limiting use to routine visits and routine clients to fit with existing norms of documentation in the ANC register, and even in one instance asking the researcher conducting the monitoring visit to assist in EDSS data entry during a busy period. The (in)coherence between the intervention-as-intended and how ANMs made sense of the intervention led to varying fidelity to point-of-care use depending on how the ANC caseload was managed. In smaller facilities with only one ANM allocated to ANC, or where only the trained ANM used the EDSS, simultaneous data entry was challenging, particularly when multiple ANC clients were waiting (negotiating capacity). ANMs snapped photos of ANC cards to input into the EDSS later, seeing participation in the EDSS intervention as simply entering the data and resulting in low fidelity to point-of-care use but higher frequency of visits entered. In facilities with moderate client caseloads yet busy ANC days, teams of ANMs worked together to deliver and document care. One ANM interacted with the ANC client and other(s) did the recordkeeping, allocating EDSS data entry alongside the paper-based forms in a parallel recordkeeping system, with both trained and untrained ANMs participating in using the EDSS. In two facilities observed using this approach, fidelity to point-of-care use remained consistently high throughout the implementation period.

Low caseloads of ANC clients hindered incorporation of the EDSS into routine practice. The EDSS software suffered technical problems, but ANMs also had few opportunities to practice using the EDSS, including for adding a follow-up visit for the same client. Several ANMs still faced skill-set workability challenges at the end of the implementation period (Table 6).

## Discussion

This study operationalised a realist assessment of implementation fidelity for an EDSS intervention to improve the quality of ANC in Nepal. We incorporated three key features: 1) we saw fidelity not as a binary (fidelity vs implementation failure) but as degrees of consistency with the intervention-as-intended; 2) we analysed fidelity as a process over time; and 3) we proposed explanations for how and why fidelity unfolded in the way that it did. We found fidelity to EDSS point-of-care use declined over time, though this fidelity component was also one of the most challenging to assess via observation. ANMs understood the intervention as primarily about recordkeeping, resulting in lower point-of-care use and often entering data after ANC visits had ended. Frequency of use for ANC visits aligned to existing paper-based recordkeeping practices, with ‘non-routine’ interactions between providers and ANC clients rarely recorded in either the EDSS or ANC registers. Quality of EDSS use (operationalised by completeness of data entry) was lower for the mIRA EDSS compared to the WHO EDSS; however, for both EDSS, and even in facilities with higher fidelity to point-of-care use, prompts and pop-ups were largely ignored.

The ‘loose’ coupling of the intervention to its implementation context enabled ANMs to negotiate use of the EDSS that at times supported fidelity (e.g., working in teams to use the EDSS at the point of care) or undermined fidelity (e.g., snapping photos of ANC cards for later EDSS data entry)(31). The taking of photos of the ANC card for later EDSS data entry was an unanticipated modification to the intervention theory that undermined the function of using the EDSS(12,14,20). Low ANC client volume at participating facilities meant that even during the monthlong supported lead-in period, ANMs had few opportunities to practice using the EDSS. Fieldworkers suggested photographing ANC cards so that ANMs could practice inputting data in the EDSS, but the suggestion almost certainly undermined the role of the EDSS to be used during the ANC consultation. It also shifted understanding of the function of the intervention from decision-aid – ideally used during interactions with clients when decisions about care were taken – to primarily a recordkeeping application. This situation demonstrates the importance of discussing the intervention theory in terms that fieldworkers and participants understand, and clarifying the intervention’s purpose and allowable adaptations so as not to undermine fidelity to its function(14).

Others have noted the difficulty in defining intervention-as-intended, and our components of fidelity presented very different implementation challenges(13). Defining fidelity in a complex intervention, like an EDSS, demands teasing out what constitutes essential elements of the intervention’s design and what constitutes the mechanisms the intervention was intended to trigger. The EDSS intervention asked healthcare providers to engage in additional recordkeeping (on top of paper-based records) and to modify how they provided ANC. Healthcare providers were expected to simultaneously provide clinical care, input information in the EDSS, and respond to its prompts. By inputting data *and* following the prompts, (we hoped) providers would deliver more guideline-recommended care components in each ANC visit. How ANMs made sense of the intervention (coherence) was critical to unpacking not just fidelity of the activity but also its intention, capturing the ‘spirit’ with which the intervention was delivered(29,32). We understood that how point-of-care use was achieved, whether via teams of ANMs or with providers working alone, was less important than fidelity to the intended function, which was use of the decision-support tool at the point where decisions about care were being made(2,8,12). However, as we found, ANMs did not view the EDSS as a decision-support tool, even when used at the point of care.

Our findings echo those from other studies examining implementation outcomes for clinical decision support tools in maternity services in low- and middle-income settings. Similar to a realist evaluation of an antenatal decision-support tool in Ghana, we found shortfalls in the skill-set workability of the intervention and uncertainty about the utility and increased workload associated with using the EDSS, particularly as in both the Ghana project and the mIRA project, the EDSS were implemented alongside paper-based recordkeeping(33). A study of an intrapartum decision-support tool in Kenya also found that providers largely used the software for recordkeeping and reporting, rather than as a clinical support tool(34).

### Limitations

Our assessment of implementation fidelity contributes to more robust evaluation and elaboration of EDSS intervention theory, offering causal explanation about what happened during implementation of the EDSS and why(9), but our study is not without limitations. Due to logistical constraints in collecting data from remote health facilities in Nepal, qualitative and quantitative data were collected at the same time, which limited probing of the fieldworkers’ observation notes based on the emerging quantitative findings. In particular, the backend data provided useful insights about unobserved EDSS usage, but as this data was not available until in-person monitoring was nearly complete, we were unable to query with ANMs about the patterns found. There were also discrepancies in the amount of data available from the difference sources for each facility. In the monitoring fieldnotes, some facilities had very detailed descriptions of implementation over time, while others had virtually no additional information documented. The monitoring fieldnotes were not wholly focused on describing implementation, with some recording (often useful) background information about staff changes and numbers of clients attending the facility or about difficulties encountered during the monitoring visit, such as not being able to observe any ANC patients or an inability to check tablet functionality (e.g., due to a forgotten password). We also found divergence between quantitative and qualitative findings that were not always easy to reconcile.

We found it challenging to identify non-mandatory fields in the backend data that could be expected to be relevant for all ANC visits in order to assess completeness of EDSS entries. The resulting non-mandatory fields assessed for the mIRA EDSS versus the WHO EDSS are qualitatively different. This limits the appropriateness of comparing quality or scope of functionality enabled between the two EDSS. The non-mandatory fields for urine protein and haemoglobin tests in the mIRA EDSS required equipment that may not have been available (though providers could indicate lack of supplies as a reason for not performing the test) and also suggested guideline-informed actions that potentially conflicted with the frequency with which ANMs thought the tests should be performed(21).

We applied Normalization Process Theory as a lens to examine the contexts and mechanisms shaping fidelity. Dalkin and colleagues have argued that Normalization Process Theory’s generative mechanisms differ ontologically from how causal mechanisms are conceived within realist evaluation, most notably that the theory’s constructs occupy the empirical realm (observable) rather than the real (unobservable)(35). While we interrogated how contexts influenced the observable actions taken by participants, we were less able to uncover the invisible drivers of action(36). Missing from this analysis were interviews with healthcare providers that could have probed the reasoning healthcare providers gave—explicitly or implicitly—for their actions and the (lack of) implementation fidelity; interviews within the longitudinal case studies were conducted prior to beginning analysis for this study.

## Conclusion

Our assessment of implementation fidelity expands the concept through a realist evaluation lens, combining a longitudinal approach to see this as a process and an emphasis on explanation of how and why patterns of implementation fidelity occurred. This leads to more nuanced understanding of the mIRA project evaluation’s result than attributing it to intervention vs implementation failure. We hoped to enhance understanding of how the EDSS intervention was enacted within the complex system of ANC in Nepal and the different kinds of work necessary to implement an EDSS in ways that enhance fidelity to its function. As clinical decision support tools continue to proliferate, more nuanced evidence on the processes driving fidelity can help improve intervention design, optimise implementation within complex systems, and enhance the potential for positive impacts on provider performance and health outcomes.

## Data Availability

All fully anonymised, quantitative data produced in the present study are available upon reasonable request to the authors. The qualitative data produced in the present study involved small numbers of participants in potentially identifiable settings that cannot be fully anonymised, so this data is not available for sharing.

## Acknowledgements

We thank Dr. Tom Fryer of University of Manchester for helpful comments on the paper’s conceptual approach. We wish to acknowledge the mIRA project team for all their work on the evaluation, including intervention implementation and the data collection that supported this study. We additionally wish to thank the healthcare providers in Nepal for their participation in the project.

## References

1. Penn-Kekana L, McPake B, Parkhurst J. Improving Maternal Health: Getting What Works To Happen. Reprod Health Matters. 2007 Jan 1;15(30):28–37.

2. Medical Research Council. Process evaluation of complex interventions [Internet]. 2015 [cited 2019 Nov 22]. Available from: https://mrc.ukri.org/documents/pdf/mrc-phsrn-process-evaluation-guidance-final/

3. Oakley A, Strange V, Bonell C, Allen E, Stephenson J. Process evaluation in randomised controlled trials of complex interventions. BMJ. 2006 Feb 16;332(7538):413–6.

4. Carroll C, Patterson M, Wood S, Booth A, Rick J, Balain S. A conceptual framework for implementation fidelity. Implement Sci. 2007 Nov 30;2(1):40.

5. Proctor E, Silmere H, Raghavan R, Hovmand P, Aarons G, Bunger A, et al. Outcomes for Implementation Research: Conceptual Distinctions, Measurement Challenges, and Research Agenda. Adm Policy Ment Health Ment Health Serv Res. 2011 Mar;38(2):65–76.

6. Dobson D, Cook TJ. Avoiding type III error in program evaluation: Results from a field experiment. Eval Program Plann. 1980 Jan 1;3(4):269–76.

7. Hasson H. Systematic evaluation of implementation fidelity of complex interventions in health and social care. Implement Sci IS. 2010 Sep 3;5:67.

8. Bonell C, Fletcher A, Morton M, Lorenc T, Moore L. Realist randomised controlled trials: A new approach to evaluating complex public health interventions. Soc Sci Med. 2012 Dec 1;75(12):2299–306.

9. Fletcher A, Jamal F, Moore G, Evans RE, Murphy S, Bonell C. Realist complex intervention science: Applying realist principles across all phases of the Medical Research Council framework for developing and evaluating complex interventions. Evaluation. 2016 Jul 1;22(3):286–303.

10. Skivington K, Matthews L, Simpson SA, Craig P, Baird J, Blazeby JM, et al. A new framework for developing and evaluating complex interventions: update of Medical Research Council guidance. BMJ. 2021 Sep 30;374:n2061.

11. Pawson, Ray, Tilley, Nick. Realistic Evaluation. London, UK: SAGE; 1997. 235 p.

12. Hawe P, Shiell A, Riley T. Complex interventions: how “out of control” can a randomised controlled trial be? BMJ. 2004 Jun 24;328(7455):1561–3.

13. Masterson-Algar P, Burton CR, Rycroft-Malone J, Sackley CM, Walker MF. Towards a programme theory for fidelity in the evaluation of complex interventions. J Eval Clin Pract. 2014;20(4):445–52.

14. Pérez D, Van der Stuyft P, Zabala M del C, Castro M, Lefèvre P. A modified theoretical framework to assess implementation fidelity of adaptive public health interventions. Implement Sci. 2016 Jul 8;11(1):91.

15. Hawe P, Shiell A, Riley T. Theorising Interventions as Events in Systems. Am J Community Psychol. 2009;43(3–4):267–76.

16. Jaspers MWM, Smeulers M, Vermeulen H, Peute LW. Effects of clinical decision-support systems on practitioner performance and patient outcomes: a synthesis of high-quality systematic review findings. J Am Med Inform Assoc JAMIA. 2011;18(3):327–34.

17. Agarwal S, Glenton C, Tamrat T, Henschke N, Maayan N, Fønhus MS, et al. Decision-support tools via mobile devices to improve quality of care in primary healthcare settings. Cochrane Database Syst Rev. 2021 Jul 27;2021(7):CD012944.

18. Mohan S, Chaudhry M, McCarthy O, Jarhyan P, Calvert C, Jindal D, et al. A cluster randomized controlled trial of an electronic decision-support system to enhance antenatal care services in pregnancy at primary healthcare level in Telangana, India: trial protocol. BMC Pregnancy Childbirth. 2023 Jan 26;23:72.

19. Haddad SM, Souza RT, Cecatti JG, Barreix M, Tamrat T, Footitt C, et al. Building a Digital Tool for the Adoption of the World Health Organization’s Antenatal Care Recommendations: Methodological Intersection of Evidence, Clinical Logic, and Digital Technology. J Med Internet Res. 2020 Oct 1;22(10):e16355.

20. Radovich E, Penn-Kekana L, Karki S, Das S, Shakya R, Campbell OMR, et al. Assessing the potential of two electronic decision support systems to improve the quality of antenatal care in primary care facilities in Nepal: study protocol [Internet]. 2023. Available from: https://figshare.com/articles/preprint/Assessing_the_potential_of_two_electronic_decision_support_systems_to_improve_the_quality_of_antenatal_care_in_primary_care_facilities_in_Nepal_study_protocol/23685099

21. Karmacharya BM, Das S, Shrestha A, Shrestha A, Karki S, Shakya R, et al. A novel approach to assessing the potential of electronic decision support systems to improve the quality of antenatal care in Nepal. Under review at Global Health: Science & Practice; 2023.

22. Jones COH, Wasunna B, Sudoi R, Githinji S, Snow RW, Zurovac D. “Even if You Know Everything You Can Forget”: Health Worker Perceptions of Mobile Phone Text-Messaging to Improve Malaria Case-Management in Kenya. PLOS ONE. 2012 Jun 13;7(6):e38636.

23. Kawamoto K, Houlihan CA, Balas EA, Lobach DF. Improving clinical practice using clinical decision support systems: a systematic review of trials to identify features critical to success. BMJ. 2005 Mar 31;330(7494):765.

24. Creswell JW, Plano Clarke VL. Designing and Conducting Mixed Methods Research. Third Edition. Thousand Oaks, California, USA: SAGE; 2018.

25. Karki S, Das S, Radovich E, Shrestha A, Shakya R, McCarthy OL, et al. The implementation realities of a digital antenatal care improvement intervention in Nepal: Auxiliary nurse midwives as street level bureaucrats. 2023.

26. Lengnick-Hall R, Gerke DR, Proctor EK, Bunger AC, Phillips RJ, Martin JK, et al. Six practical recommendations for improved implementation outcomes reporting. Implement Sci IS. 2022 Feb 8;17:16.

27. Brooks J, McCluskey S, Turley E, King N. The Utility of Template Analysis in Qualitative Psychology Research. Qual Res Psychol. 2015 Apr 3;12(2):202–22.

28. Fryer T. A critical realist approach to thematic analysis: producing causal explanations. J Crit Realism. 2022 Aug 8;21(4):365–84.

29. May CR, Albers B, Bracher M, Finch TL, Gilbert A, Girling M, et al. Translational framework for implementation evaluation and research: a normalisation process theory coding manual for qualitative research and instrument development. Implement Sci. 2022 Feb 22;17(1):19.

30. May C, Finch T. Implementing, Embedding, and Integrating Practices: An Outline of Normalization Process Theory. Sociology. 2009 Jun 1;43(3):535–54.

31. May CR, Johnson M, Finch T. Implementation, context and complexity. Implement Sci. 2016 Oct 19;11(1):141.

32. Steckler A, Linnan L, editors. Process Evaluation for Public Health Interventions and Research. Jossey-Bass/Wiley; 2002.

33. Abejirinde IOO, Zweekhorst M, Bardají A, Abugnaba-Abanga R, Apentibadek N, Brouwere VD, et al. Unveiling the Black Box of Diagnostic and Clinical Decision Support Systems for Antenatal Care: Realist Evaluation. JMIR MHealth UHealth. 2018;6(12):e11468.

34. Dinh N, Agarwal S, Avery L, Ponnappan P, Chelangat J, Amendola P, et al. Implementation Outcomes Assessment of a Digital Clinical Support Tool for Intrapartum Care in Rural Kenya: Observational Analysis. JMIR Form Res. 2022 Jun 20;6(6):e34741.

35. Dalkin SM, Hardwick RJL, Haighton CA, Finch TL. Combining Realist approaches and Normalization Process Theory to understand implementation: a systematic review. Implement Sci Commun. 2021 Jun 26;2(1):68.

36. Dalkin SM, Greenhalgh J, Jones D, Cunningham B, Lhussier M. What’s in a mechanism? Development of a key concept in realist evaluation. Implement Sci. 2015 Apr 16;10(1):49.

